# Sexual risk among pregnant women at risk of HIV infection in Cape Town, South Africa: What does alcohol have to do with it?

**DOI:** 10.1101/2021.11.30.21267089

**Authors:** Amanda P. Miller, Steven Shoptaw, Rufaro Mvududu, Nyiko Mashele, Thomas J. Coates, Linda-Gail Bekker, Zaynab Essack, Candice Groenewald, Zaino Peterson, Pamina M. Gorbach, Landon Myer, Dvora L. Joseph Davey

## Abstract

This study examines associations between alcohol use and HIV sexual risk among a cohort of HIV-uninfected pregnant women (n=1201) residing in a high HIV burden community in South Africa. Alcohol use was measured using a modified version of the Alcohol Use Disorder Identification Test (AUDIT). HIV sexual risk was measured through a composite variable of four risk factors: diagnosis with a STI, self-report of >1 recent sex partners, partner HIV serostatus (unknown or HIV+) and condomless sex at last sex. Any past year alcohol use prior to pregnancy was reported by half of participants (50%); 6.0% reported alcohol use during pregnancy. Alcohol use prior to pregnancy was associated with increased odds of being at high risk of HIV (aOR=1.33 for 2 risks and aOR=1.47 for 3 risks). In addition to reducing alcohol use, several other strategies to address HIV sexual risk in this population were identified.

## Introduction

Heavy alcohol use and HIV are prevalent interrelated public health issues that are associated with a substantial burden of disease in South Africa (1). With a national HIV prevalence of 19%, South Africa is home to the most persons living with HIV globally (7.8 million) (2). Although only 31% of South Africa’s population reports alcohol use in the past 12 months, consumption levels are high among those who drink, with 59% engaging in heavy episodic drinking (HED), which is defined as the consumption of 60 grams or more of pure alcohol (the equivalent of six standard drinks) in one sitting (3). Estimates for total per capita consumption among drinkers (29.9 liters of pure alcohol), prevalence of alcohol use disorders (AUD) (7%) and prevalence of alcohol dependence (2%) in South Africa exceed regional averages (3). There are gendered differences in patterns of use and quantity of alcohol consumption in South Africa, with men being both more likely to drink and to consume hazardous levels of alcohol (3). However, HED is prevalent among women who consume alcohol. A recent report by World Health Organization (WHO) found that roughly one-third (34%) of women reported having engaged in this behavior in the month preceding the survey (3) while another study found that a similar proportion of women (32%) reportedly engage in binge drinking on a weekly basis (4). Alcohol use is also common among pregnant women in Cape Town (5–8). Prior work among pregnant women in this setting found that 29% reported alcohol use during pregnancy (9), an estimate that is likely biased due to underreporting/social desirability bias (10). Biomarker measures of hazardous alcohol use (23%) have also confirmed use among pregnant women who are living with HIV in South Africa (11).

Both alcohol use and incident HIV infection during pregnancy are associated with adverse health outcomes for the mother and fetus. Fetal alcohol spectrum disorder (FASD) describes a continuum of cognitive and physical impairments a fetus may experience due to prenatal alcohol exposure (PAE), the most severe form being fetal alcohol syndrome (FAS) (12). As a result of heavy alcohol use in pregnancy, South Africa has the highest rate of FAS in the world (13). FAS is a lifelong diagnosis and its deleterious effects on the central nervous system can result in neurocognitive developmental delays and behavioral problems that can persist into adolescence and young adulthood (14, 15). There is also evidence of socioeconomic disparities in the “harm per liter” of alcohol use, with poor South Africans experiencing worse outcomes associated with their alcohol use. Sixty percent of deaths attributable to alcohol use occur among persons in the bottom third of the socioeconomic spectrum (16). These socioeconomic disparities also extend to alcohol attributable HIV mortality (17). In addition to directly threatening the health of both mother and baby, alcohol use is a widely recognized driver of the HIV epidemic.

Alcohol use can impact decision making around sexual practices including having multiple sex partners (18, 19) and condomless sex (18, 20), resulting in increased risk of HIV acquisition (21–24). Alcohol use is also associated with increased risk of intimate partner violence (IPV) (25, 26), which itself is a risk factor for HIV, through forced sex and reduced ability to safely negotiate condom use (27). While the evidence base supporting alcohol use as a risk factor for engaging in sexual risk behavior is well established (21), data examining this relationship among pregnant women in South Africa is more limited. Prior work with pregnant women found that alcohol use prior to (Cape Town (28)) or during (Mpumalanga (29)) pregnancy was associated with having multiple sexual partners but not condomless sex (29). In another Cape Town study one in four pregnant women (27%) indicated that alcohol and drug use leads to greater sexual risk taking (30). Alcohol use is also associated with barriers to optimum health outcomes in the pre-exposure prophylaxis (PrEP) care continuum, around initiation and adherence (31), reducing the utility of the most effective biomedical intervention available to reduce HIV acquisition in individuals at high risk of infection. A robust body of literature suggests that alcohol use is also a barrier to ART uptake and adherence among persons living with HIV (32), including pregnant women (33). The effects of alcohol use on HIV risk, including non-adherence to PrEP, are especially problematic in pregnant women, due to elevated risk for HIV acquisition during pregnancy (34) as well as increased risk of vertical transmission due to high maternal viral load during the acute infection phase (35, 36). These intertwined public health issues of alcohol use, risky sexual practices, IPV and HIV can result in excess disease burden and exacerbate health disparities.

Addressing alcohol use among pregnant women is critical to HIV prevention efforts and public health efforts to promote healthy pregnancies and births. Given that alcohol use is a modifiable behavior, the importance of characterizing alcohol use, and identifying associations between alcohol and other HIV risk factors among pregnant women who experience a high burden of HIV incidence cannot be overlooked. This knowledge can inform the development of contextually sensitive interventions to interrupt and potentially prevent alcohol use during pregnancy. Given that withdrawal can be medically dangerous among persons experiencing alcohol dependence (for both mother and fetus) it is also important to identify women experiencing alcohol dependence in pregnancy to refer them to treatment centers that can facilitate supervised detoxification (37). The present analysis fills an important gap in the existing literature regarding alcohol use as a driver of sexual risk behavior among pregnant women by exploring associations between alcohol use and HIV sexual risk, among a large cohort of pregnant women residing in an underserved community experiencing a heavy burden of HIV in Cape Town, South Africa. We measure HIV sexual risk using a composite proxy risk measure to identify modifiable risk factors and hypothesize that alcohol use will be associated with greater odds of being in the high-risk category. We also examine sociodemographic and behavioral factors associated with alcohol use in this population.

## Methods and Materials

### Participants and Procedures

Pre-exposure prophylaxis in Pregnancy and Post-partum (PrEP-PP) is an ongoing observational prospective closed-cohort study of 1201 pregnant and postpartum women attending antenatal care (ANC) in Gugulethu, Cape Town, South Africa (clinical trial no. NCT03902418). This community was selected because of the high incidence and prevalence of HIV among pregnant and breastfeeding women (38). Consecutive eligible, consenting study participants were enrolled at the study site until the target sample size of 1201 pregnant women was met. Women 16 years or older attending their first ANC visit and pregnant at time of enrollment were eligible for study participation. Additional inclusion criteria included confirmed HIV negative serostatus (confirmed with 4^th^ generation ab/ag Abbott rapid test), intention to give birth in Cape Town, no medical or psychiatric conditions contraindicated for PrEP and consent to participate in the study. Participants are followed until 12 months’ post-partum or until censorship (pregnancy loss, infant death, seroconversion, moving away, transfer out of care, loss to follow-up). Consented, enrolled participants are invited to return every 3 months for study visits that correspond with their ANC visits. All study staff are trained, salaried staff working for University of Cape Town.

The present analysis utilizes baseline data (n=1,201) from study enrollment visits, collected between August 2019 and September 2021. Trained research staff conducted eligibility screening, HIV counseling and testing, counseling around HIV prevention and PrEP use, offer to initiate PrEP, diagnostic testing for other STIs and a counselor administered survey which takes approximately 30-45 minutes to complete. For STI testing and management, participants were instructed on how to do a self-collected vaginal swab which was tested for Chlamydia trachomatis (CT), Neisseria gonorrhoeae (NG), and Trichomonas vaginalis (TV) using point of care testing (Cephid, Inc., Sunnyvale, CA, US). Treatment is provided as needed during the same visit following South Africa National STI Guidelines (39). The study interviewers collect data on participant sociodemographics, sexual practices, depression, alcohol use, and attitudes and knowledge towards PrEP.

Written informed consent was obtained from all study participants; unassisted self-consent was obtained from adolescents (participants 16 and 17 years of age). The study received approval from UCT Human Research Ethics Committee (HREC) to waive parental consent. Participants were given the informed consent form to take home if they wanted to discuss with partners or parents and could return within 24 hours if they wanted to participate in the study. Participants viewed a video describing the study and the consent process in isiXhosa, the primary language spoken in this setting, to ensure that they understood the study design and consent form fully before signing. Light refreshments, compensation for their time (R100, ∼USD$7) as well as reimbursement for transportation are provided at each study visit (40). This study was approved by University of Cape Town Faculty of Health Sciences Human Research Ethics Committee (UCT-HREC) and University of California Los Angeles (UCLA).

### Measures

Alcohol use, our exposure of interest, was measured using a modified versions of the 10 item Alcohol Use Disorders Identification Test (AUDIT) (41, 42), a validated measure that has been used globally, including among pregnant women in Cape Town (43, 44). The AUDIT typically asks questions for the reference period of “the past year”. For the present study, questions were modified to the reference period “since finding out you were pregnant”. The AUDIT is scored by summing responses to the 10 items (score range of 0-40 with high scores indicative of greater alcohol use) and a cutoff is used to differentiate between lower risk use, hazardous or harmful use and persons with probable alcohol dependence. A subset of the AUDIT, the first three AUDIT questions which are used to measure alcohol consumption levels (AUDIT-C (45)), were used to identify hazardous alcohol use in the year prior to pregnancy (range 0-12). AUDIT-C questions were also asked for the reference period, “in the past year prior to finding out you were pregnant”. Given the adverse consequences of any amount of alcohol use while pregnant on neonatal outcomes, the potential for underreporting of alcohol use while pregnant, and prior work which suggests that alcohol use prior to pregnancy is a good predictor of alcohol use during pregnancy (46), our primary alcohol exposure of interest was any alcohol use prior to pregnancy (an AUDIT-C score of 0). Limited data on patterns of alcohol use in South Africa immediately prior to and during pregnancy exist; to address this gap, we also explored patterns of risky alcohol use prior to and during pregnancy as secondary exposures of interest. We consider all alcohol use during pregnancy to be “risky”; therefore, we adopted a conservative AUDIT cutoff of ≥5 to capture even light to moderate patterns alcohol use which we would consider “higher risk” during pregnancy. This “higher risk” cutoff captures the categories of risky, harmful, and severe alcohol use in women and has previously been used among pregnant women in South Africa (43). To identify women with probable alcohol dependence, we used a cut-off of ≥20. For the AUDIT-C we used a cut-off of ≥3 for hazardous drinking, a cut-off previously used with pregnant women in South Africa (11).

HIV risk, our primary dependent variables of interest, was measured through a proxy composite variable of four risk factors: (1) diagnosis with a STI infection (measured at baseline), (2) self-report of multiple sexual partners (reporting more than one partner in the past 3 months), (3) self-report of partner HIV serostatus (dichotomized into two categories: HIV-negative/no partner and HIV-positive/unknown serostatus), and (4) self-report of condomless sex at last sex. An ordinal count variable was created for individuals experiencing these risk factors. A sensitivity analysis was performed to identity an optimum cutoff and a binary composite HIV sexual risk variable was created for those at high-risk vs not high-risk. Results are presented for two cut offs: we explore classifying women who experience two or more HIV risk factors as high risk as well as classifying women who experience three or more HIV risk factors as high risk.

Other measures analyzed in descriptive and multivariable analysis included sociodemographic variables such as age, educational attainment (did not complete secondary school/completed secondary school), employment status (a six category variable collapsed into a three category variable: any form of employment/unemployed/student), residence type (a five category variable collapsed into a binary variable: formal/informal housing) and beliefs that may impact alcohol use during pregnancy such as participants’ pregnancy intentions, which were measured using items adapted from prior studies of fertility intentions in this setting (47, 48), including feelings about the timing of the pregnancy (did this pregnancy happen at the right/wrong time?) and having a baby (I wanted to have a baby/I have mixed feelings about having a baby/I did not want to have a baby). HIV risk factors were also considered, including current sexual partner (no/yes, the father of my unborn baby/yes someone else), relationship status (cohabitating/not cohabitating), likelihood their partner has other sexual partners (not likely at all/ somewhat likely/very likely/I don’t know) and frequency of condom use in past 3 months (never/rarely/sometimes/almost always/always). Other risk factors included depression, using the Edenborough Post-natal Depression Scale (EPDS) (49) (cutoff of ≥11 for depression (50)) and experiences of past year verbal, physical and sexual IPV using 13 items adapted from the WHO IPV Scale (collapsed into a binary variable: any/no recent experiences of IPV) (51, 52).

### Data Analysis

Analyses were conducted using SAS studio (53). The data were first inspected for errors, omissions, and values lying outside of the limit ranges. In order to identify the most appropriate alcohol use measure to use in the present analysis, a sensitivity analysis was performed. The analytic sample was restricted to participants who provided responses to our primary exposure of interest, alcohol use, resulting in an analytic sample of 1201. The distribution, mean and median, for each of our alcohol measures were explored as well as the internal reliability (Cronbach’s alpha coefficient) and cut-off points were informed by the existing literature. Prior research has found that alcohol use prior to pregnancy is a good predictor of alcohol use in pregnancy (46). It is also worth noting that prior work in this study sample found a median gestational age at baseline (first ANC visit) of 21 weeks (the fifth month of pregnancy) (54), so “prior to pregnancy” may encompass the entire first trimester, a critical time in fetal development. Based on this sensitivity analysis and our prior work it was determined that past year alcohol use prior to pregnancy would be the alcohol measure used in bivariate and multivariable analyses. Next, sociodemographic and HIV risk related behavioral variables of interest were analyzed, using descriptive statistics, to characterize the analytic sample, overall, and by alcohol use. Stratified bivariate analysis of covariates by any past year alcohol use prior to pregnancy was performed using χ2 analysis, fisher’s exact test and two sample t-test. Statistical significance was determined using an alpha of 0.05.

To test our two hypotheses, multivariable logistic regression models were built using the proc logistic function and the logit link to explore associations between past year alcohol use prior to pregnancy and our HIV risk outcome. Directed acyclic graphs were used *a priori* to determine the minimally sufficient set of covariates to adjust for to reduce bias from measured confounders (see supplemental file). A variable was considered a confounder if it was predictive of both the exposure and outcome and was not on the causal pathway between exposure and outcome. Multicollinearity was assessed by examining the intercorrelations between the exposure variables in the model as well as the tolerance and variance inflation factor (VIF). All VIF approximated 1, no tolerance values were <0.1 and no correlations exceeded 0.7.

## Results

### Description of the study sample

Table 1 provides relevant sociodemographic and behavioral characteristics from baseline visits (i.e., first ANC appointment) for the 1201 study participants. Characteristics are provided for the overall sample and stratified by past year alcohol use prior to pregnancy. Mean age of participants was 26.6 years (SD 5.9); roughly half (51%, n=617) had completed secondary schooling and resided in an informal dwelling (54%, n=644). The median gestational age was 21 weeks (IQR 15-31). Half (50%, n=598) felt the timing of the pregnancy was wrong and nearly half (49%, n=585) reported they did not want to have the baby. Past year IPV was reported by 12% (n=147) of participants and 7% (n=89) of participants had EPDS scores indicative of current depression. Most participants currently had a sexual partner (92%, n=1,105) and this partner was almost always the baby’s father. Nearly one third of participants (29%, n=345) were diagnosed with a STI infection; more than one in five participants (22%, n=268) were unaware of their partners HIV serostatus while 2% (n=18) reporting having a partner living with HIV. Most participants (69%, n=804) reported never using a condom in the prior three months. Mean age, residence type, past year IPV, having multiple sexual partners, pregnancy timing, feelings about the pregnancy, cohabitation and primary partner’s HIV status significantly differed by alcohol use status. Persons reporting alcohol use (versus no alcohol use) were more likely to be younger (mean age: 25.9 years vs 27.3 years), less likely to feel the pregnancy timing was good (45% vs 55%), less likely to report wanting to have the baby (30% vs 42%), more likely to have a partner of unknown HIV serostatus (25% vs 19%), more likely to report IPV (17% vs 8%) and more likely to have multiple partners (5% vs 1%).

**Table 1.**
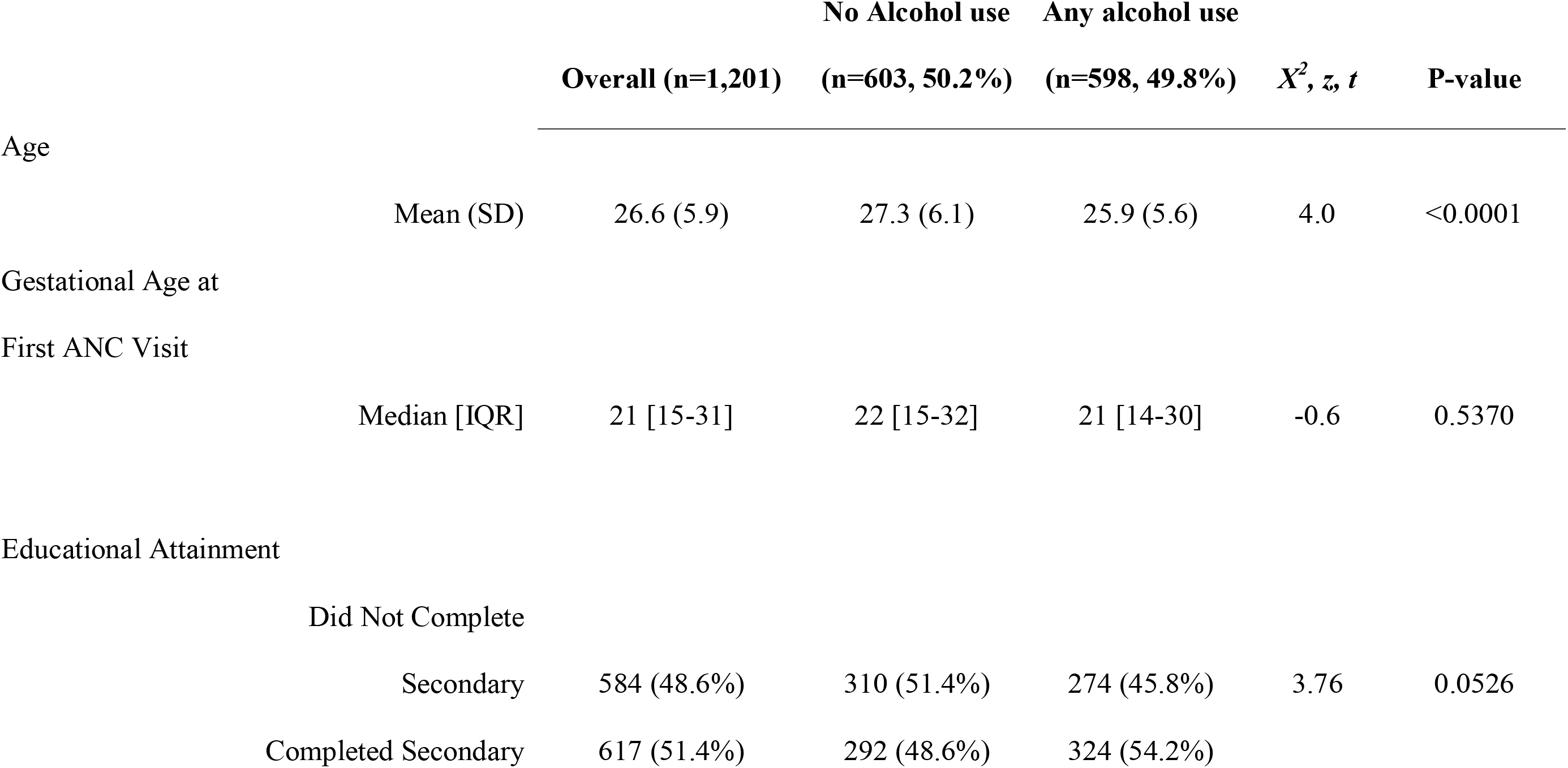
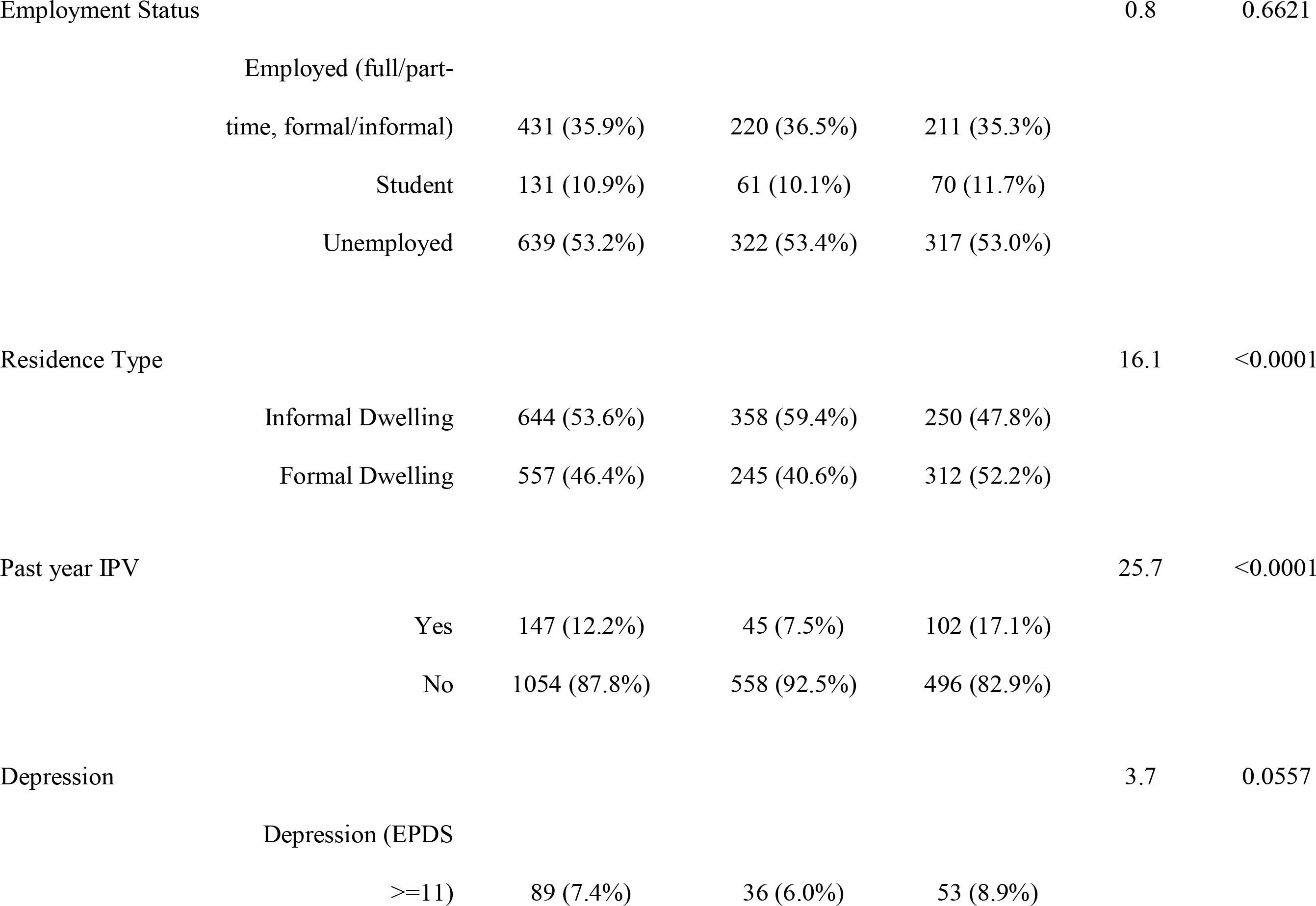
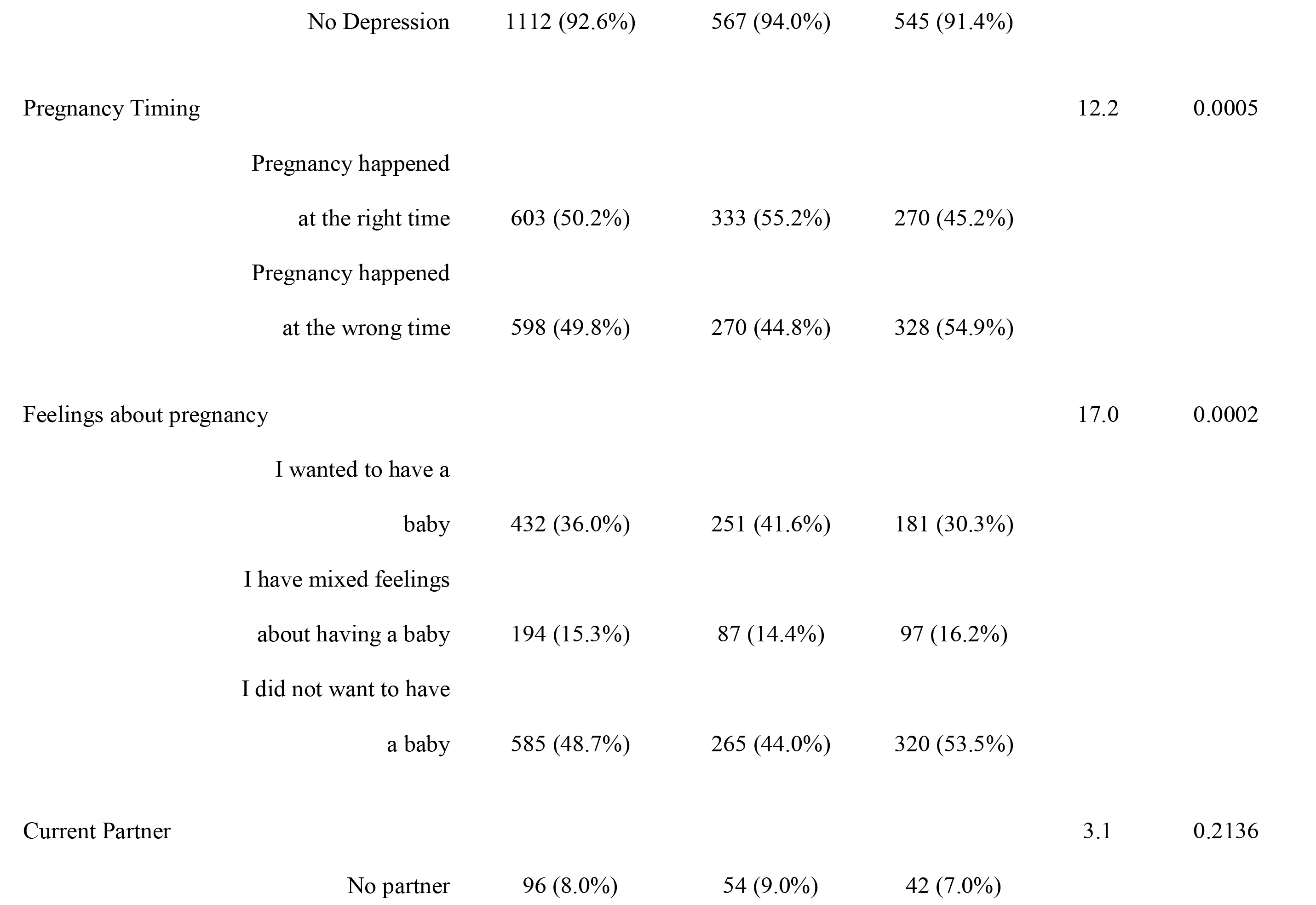
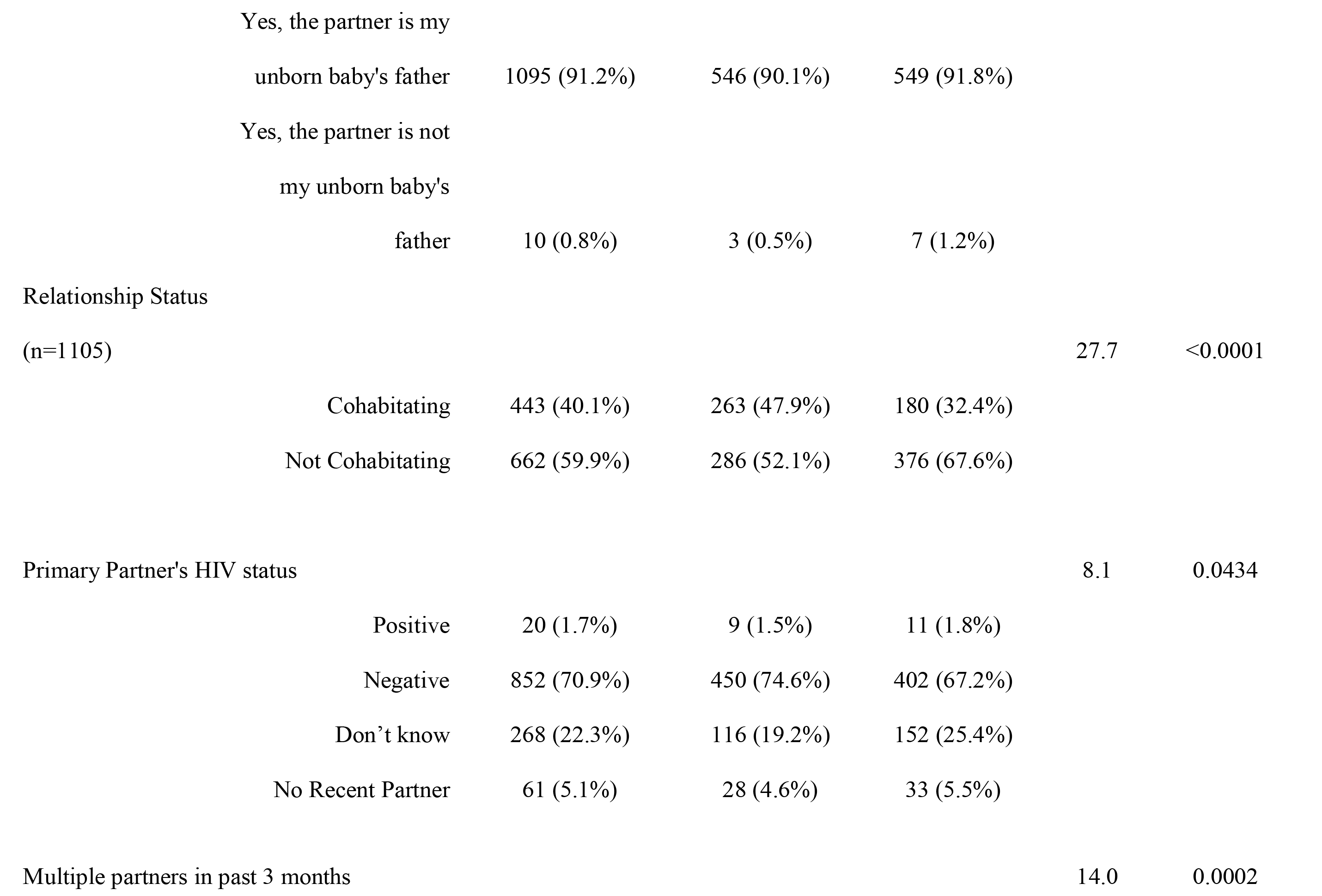
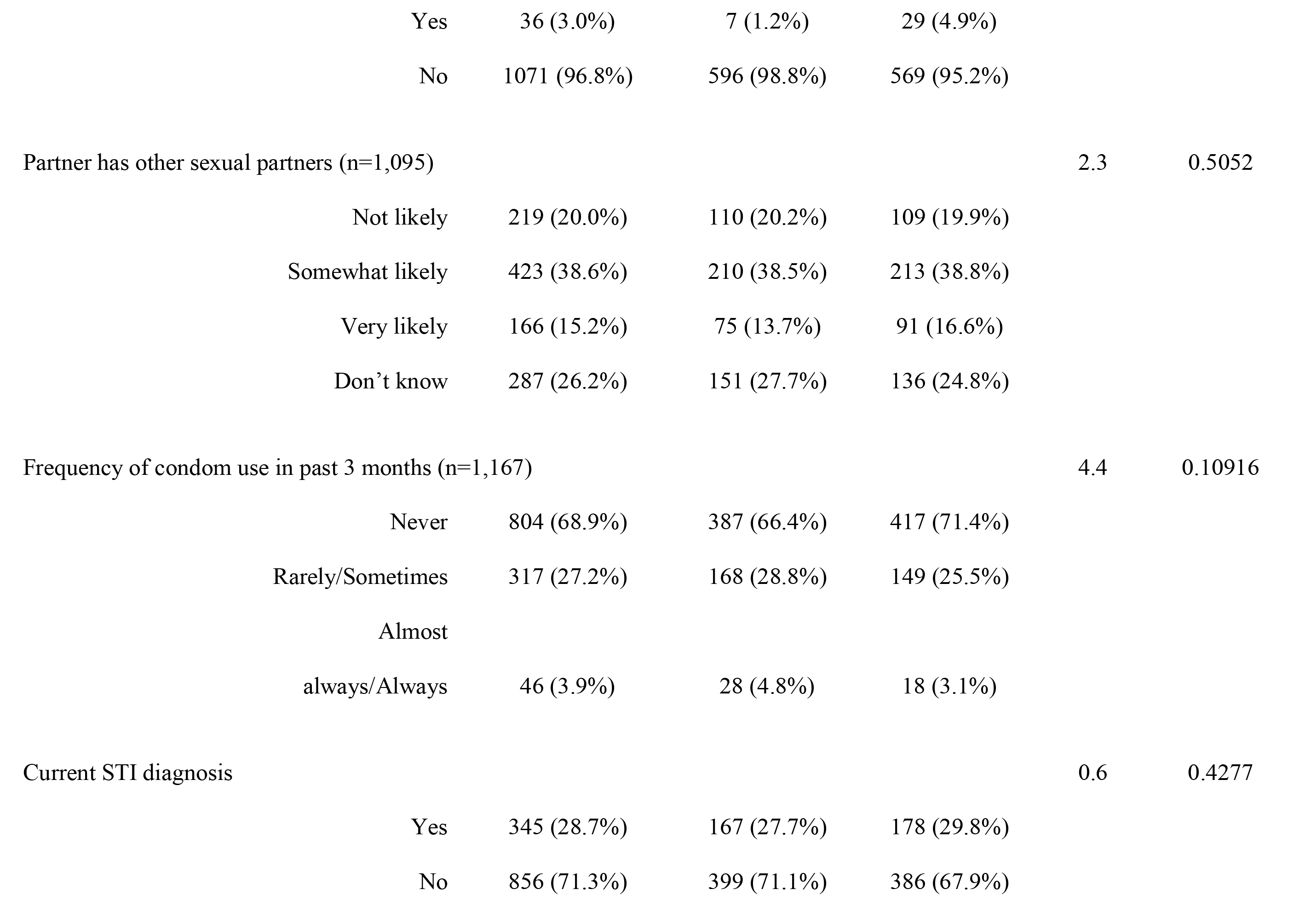
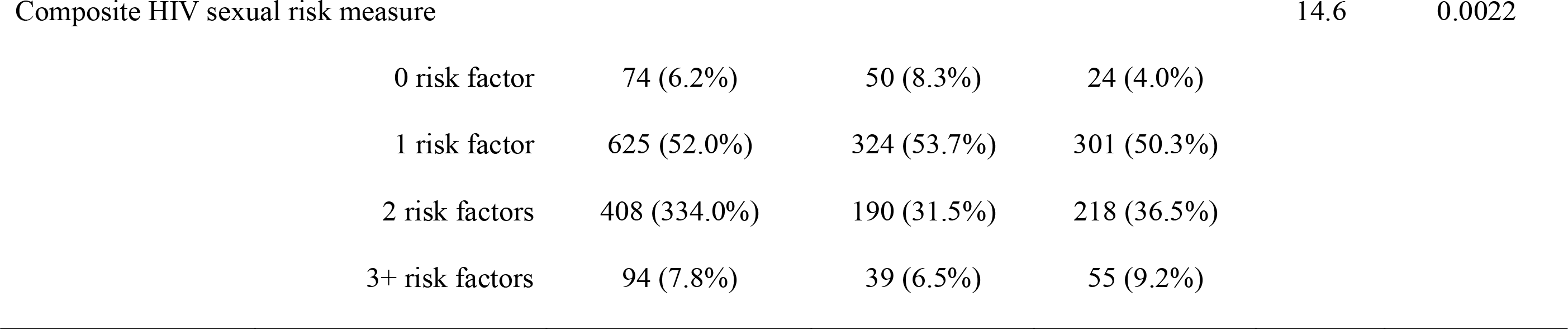
Sociodemographic and behavioral associated with alcohol use in 12 months prior to pregnancy among pregnant women at risk of HIV in Cape Town, South Africa (n=1201 women)

### Sensitivity analysis of alcohol use measures in the study sample

Results from the sensitivity analysis of our alcohol use measures before and during pregnancy can be found in Table 2. Any alcohol use in the past year prior to pregnancy was reported by half of participants (50%, n=598). Prior to pregnancy, one-third (33%, n=396) of the sample engaged in hazardous drinking; when looking exclusively among those reporting any alcohol use during this time frame, the majority (66%, n=396) engaged in hazardous drinking. Six percent of participants reported alcohol use during pregnancy. Of those reporting any alcohol use during this time frame, using a conservative AUDIT cutoff to capture the categories of risky, harmful and severe alcohol use, 46% (n=33) of pregnant women reporting any alcohol use during pregnancy had AUDIT scores indicative of higher risk alcohol use. One third (32%, n=23) of pregnant women reporting any alcohol use during pregnancy had AUDIT-C scores indicative of heavy alcohol use. Three percent of women reporting any alcohol use during pregnancy had an AUDIT score indictive of probable alcohol dependence. The AUDIT and AUDIT-C (for both reference periods: during and before pregnancy) had Cronbach’s alpha coefficients suggestive of good internal consistency (>0.8) in our study sample.

**Table 2.**
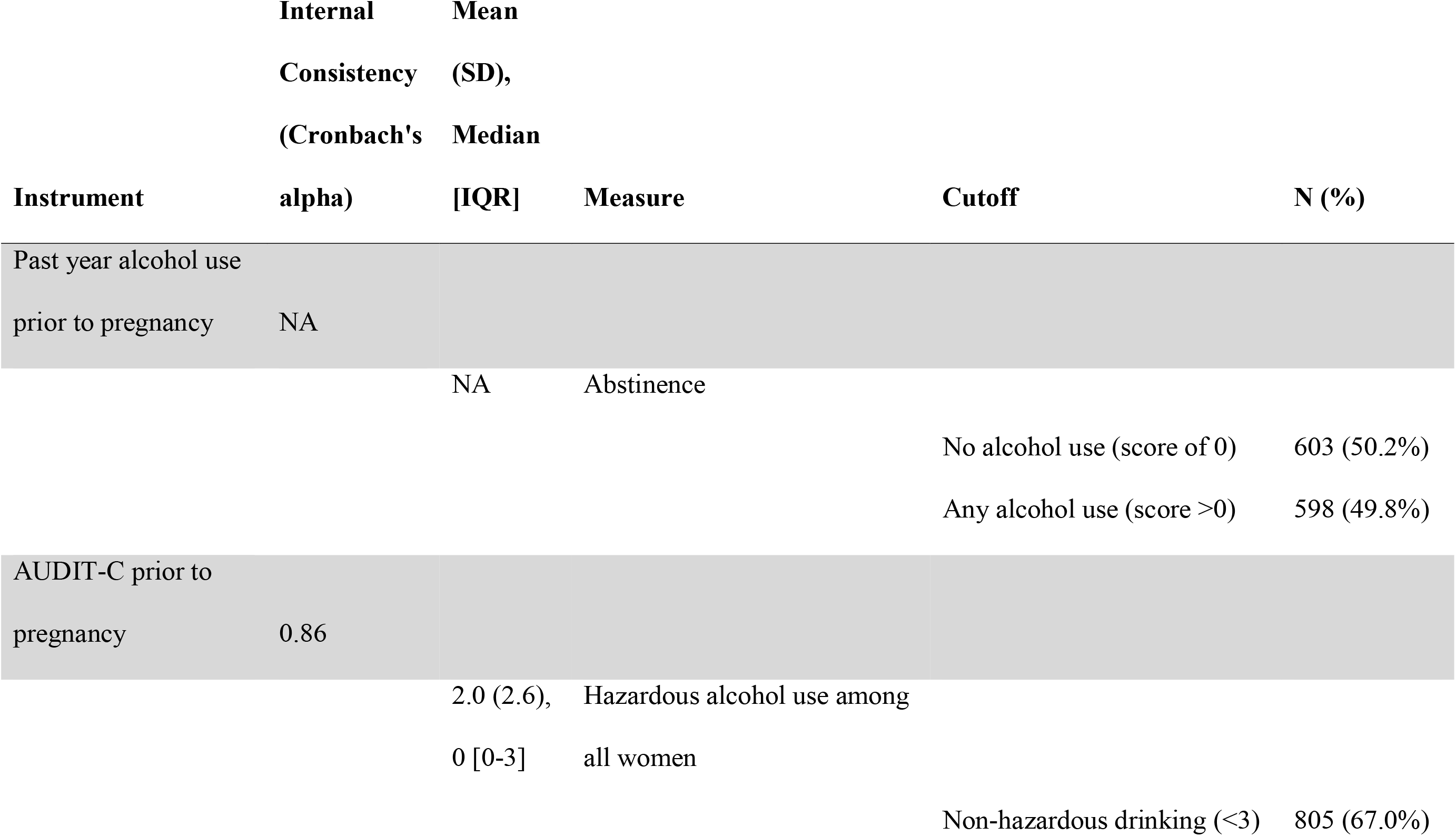
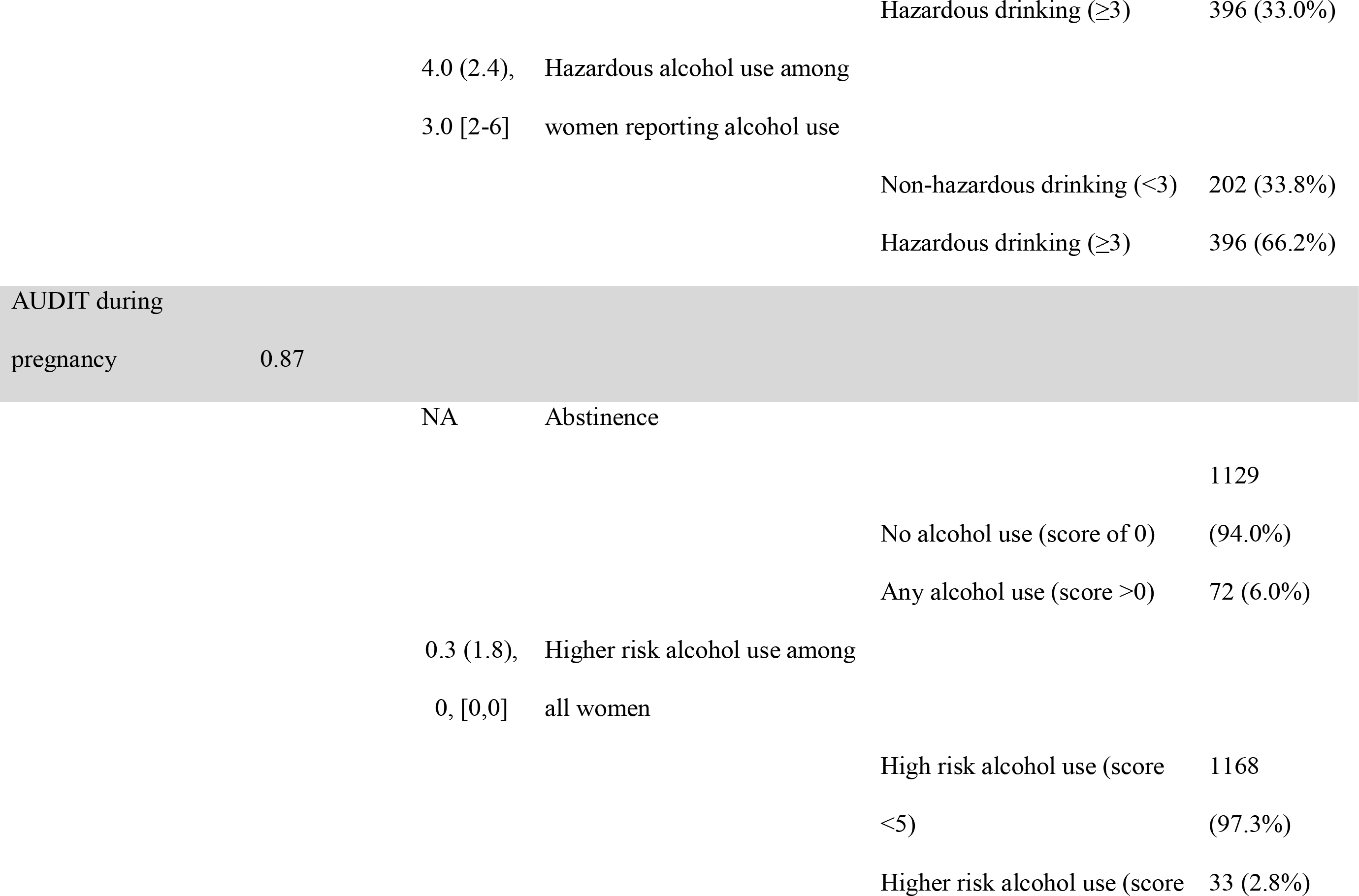
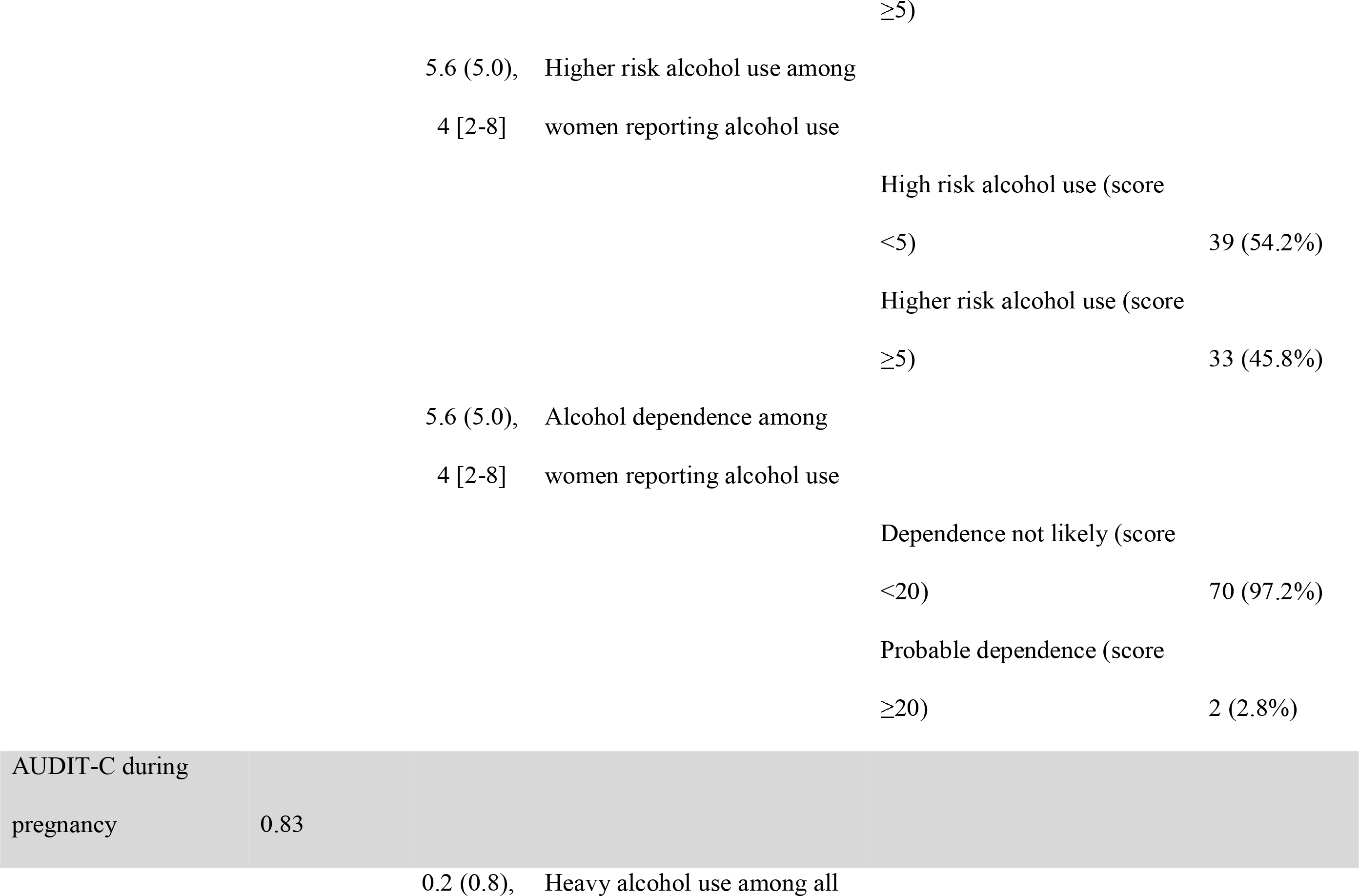
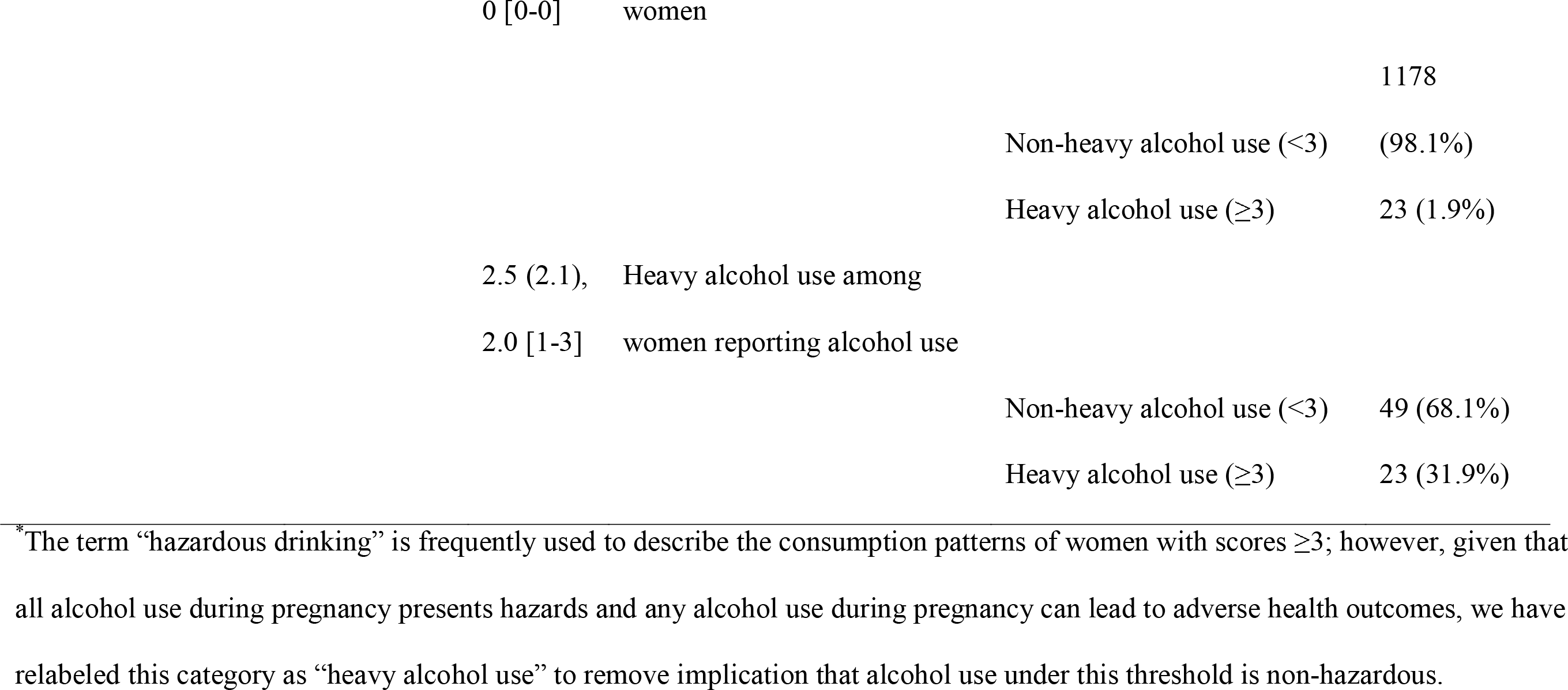
Performance of Alcohol Use Measures in the Study Sample Among Pregnant Women in Cape Town, South Africa (n=1,201)

### Descriptive characteristics of women at highest risk of HIV

Table 3 presents sociodemographic characteristics of women in the highest risk factor (3+ risks) group (n=94). Among women with three of more HIV risks factors, mean age of participants was 25.8 years (SD 6.3); 38% (n=36) had completed secondary schooling and 62% (n=58) resided in an informal dwelling. Fifty nine percent (n=55) reported alcohol use prior to pregnancy while 9% (n=8) reported alcohol use during pregnancy. The majority (58%, n=54) felt the timing of the pregnancy was wrong and more than half (56%, n=53) reported they did not want to have the baby. Past year IPV was reported by 14% (n=13) of participants and 5% (n= 5) of participants had EPDS scores indicative of depression. Nearly all participants currently had a sexual partner (99%, n=93) and this partner was almost always the baby’s father. The most common risk factor was condomless sex at last sex, reported by 99% (n=92) of participants, followed by a partner of positive or unknown HIV serostatus (97%, n=91), current STI infection (93%, n=87) and 15% (n=14) had multiple sexual partners in the past 3 months. Three percent of women (n=3) in this high-risk category had all four HIV risk factors.

**Table 3.**
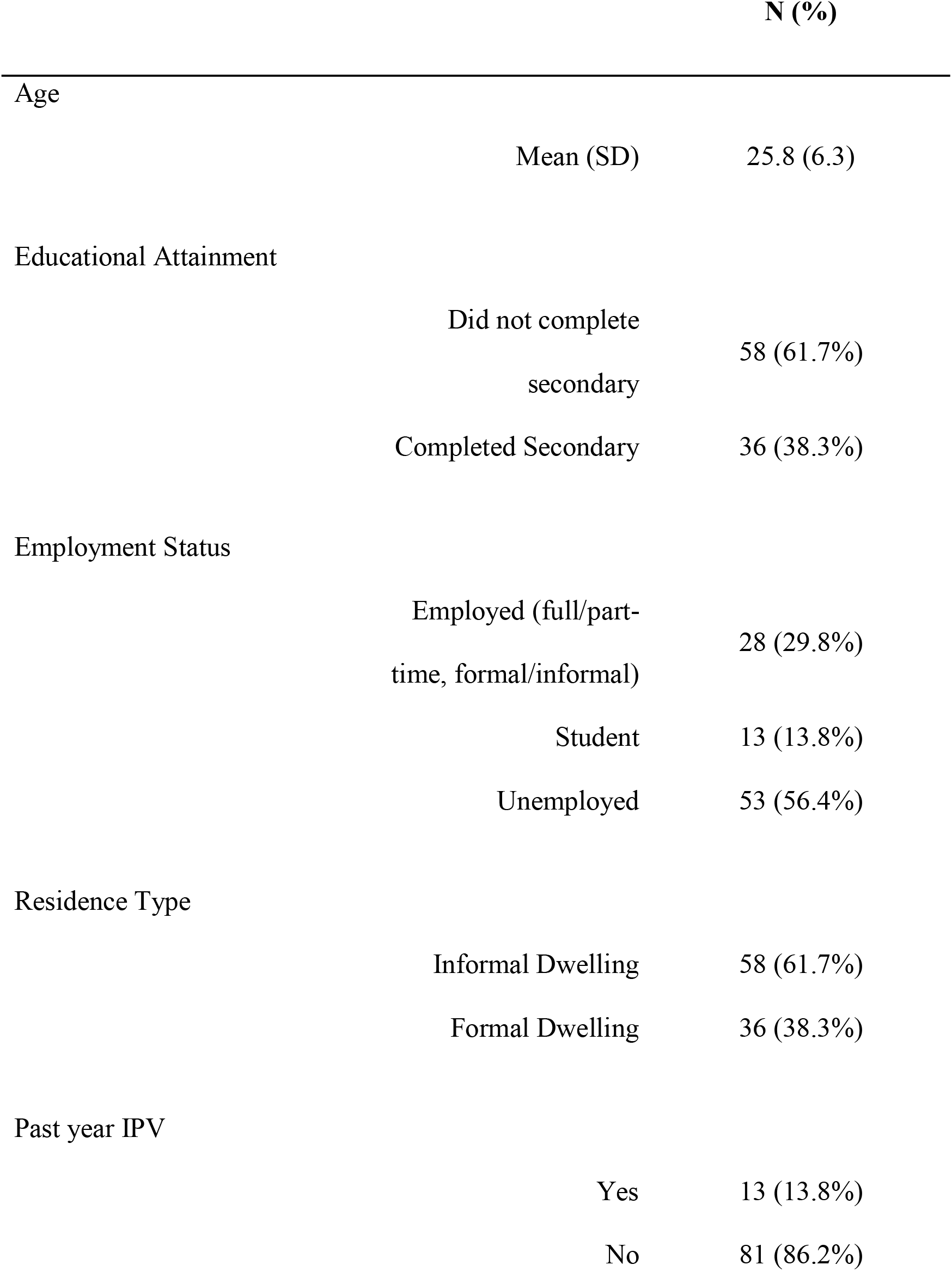
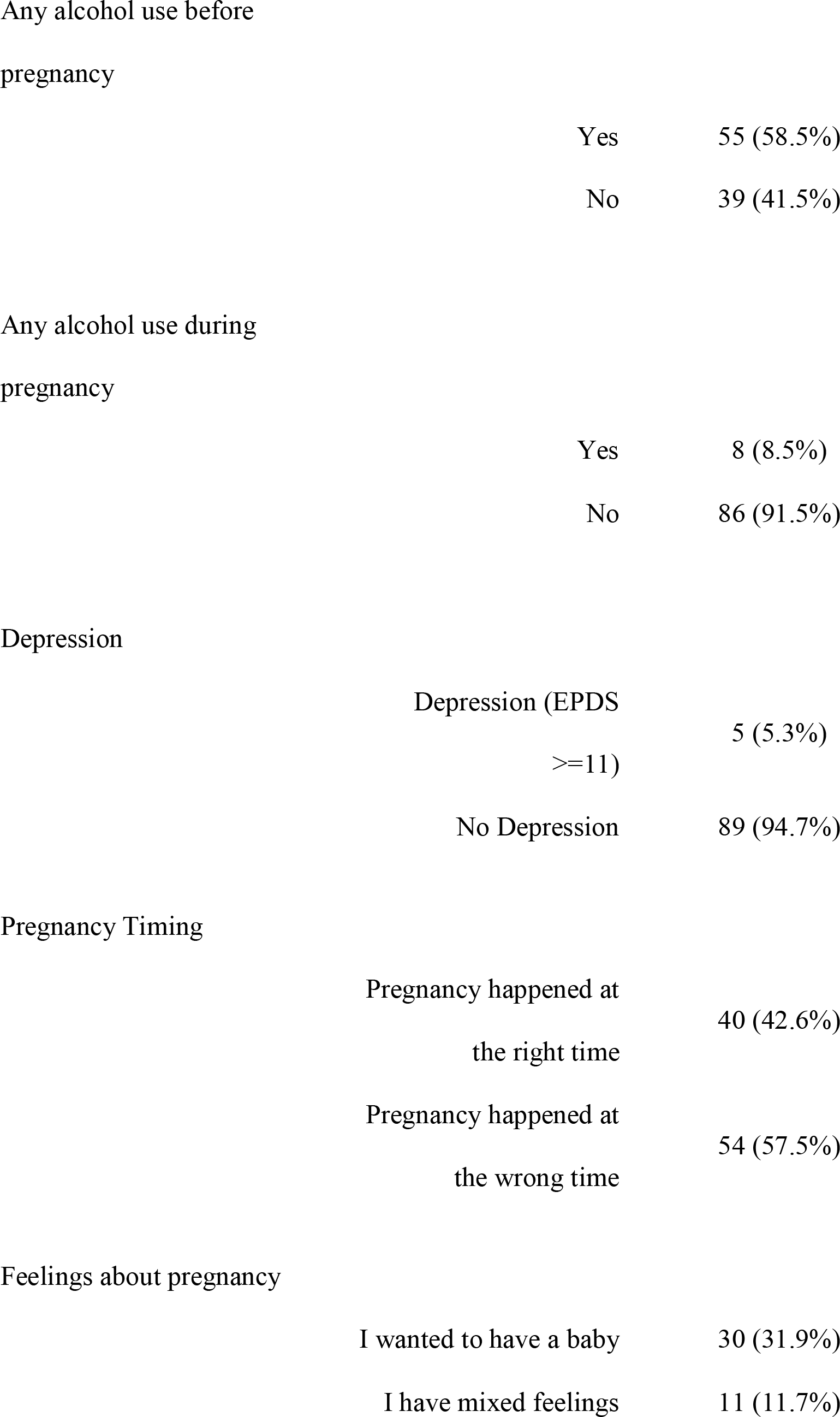
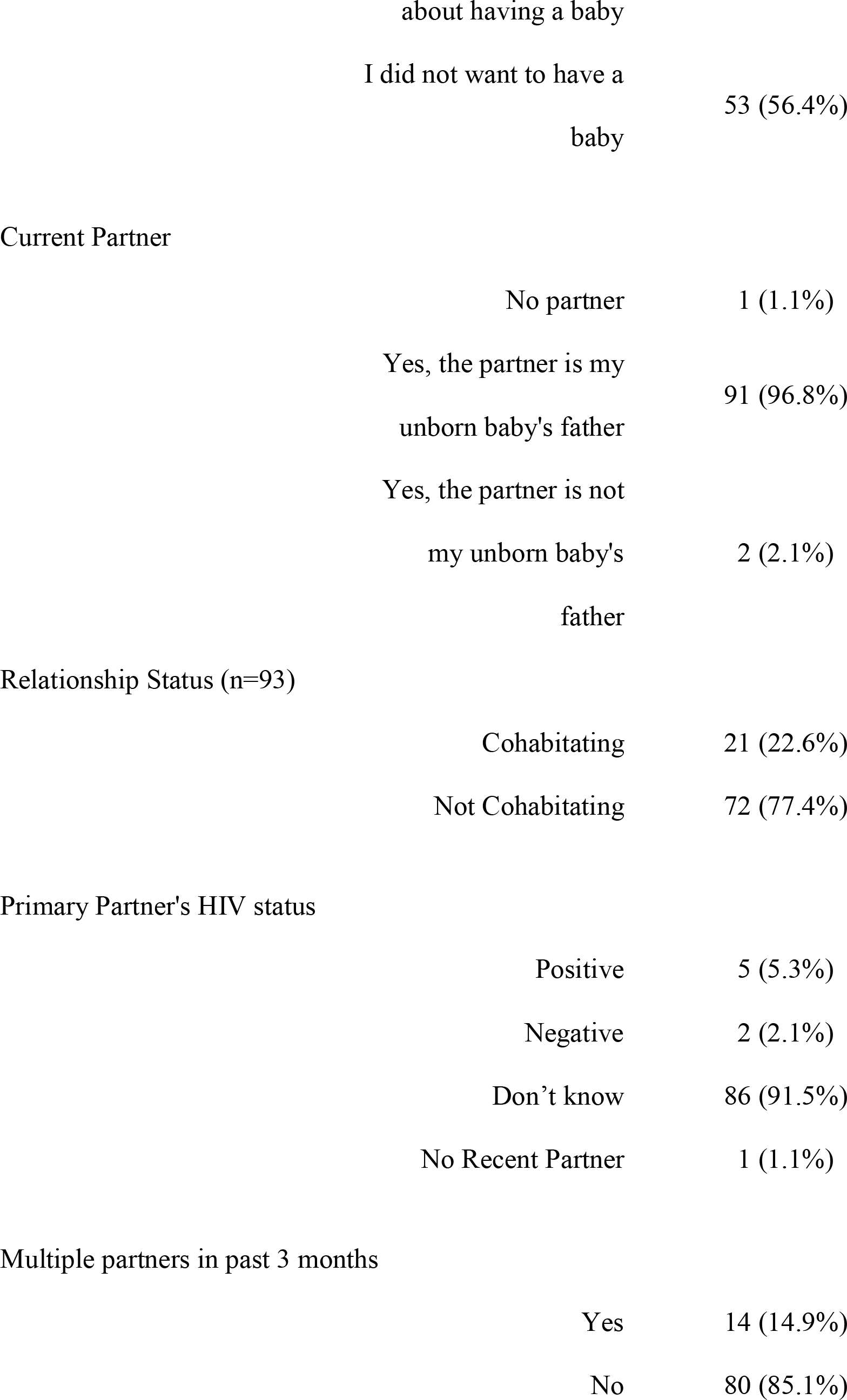
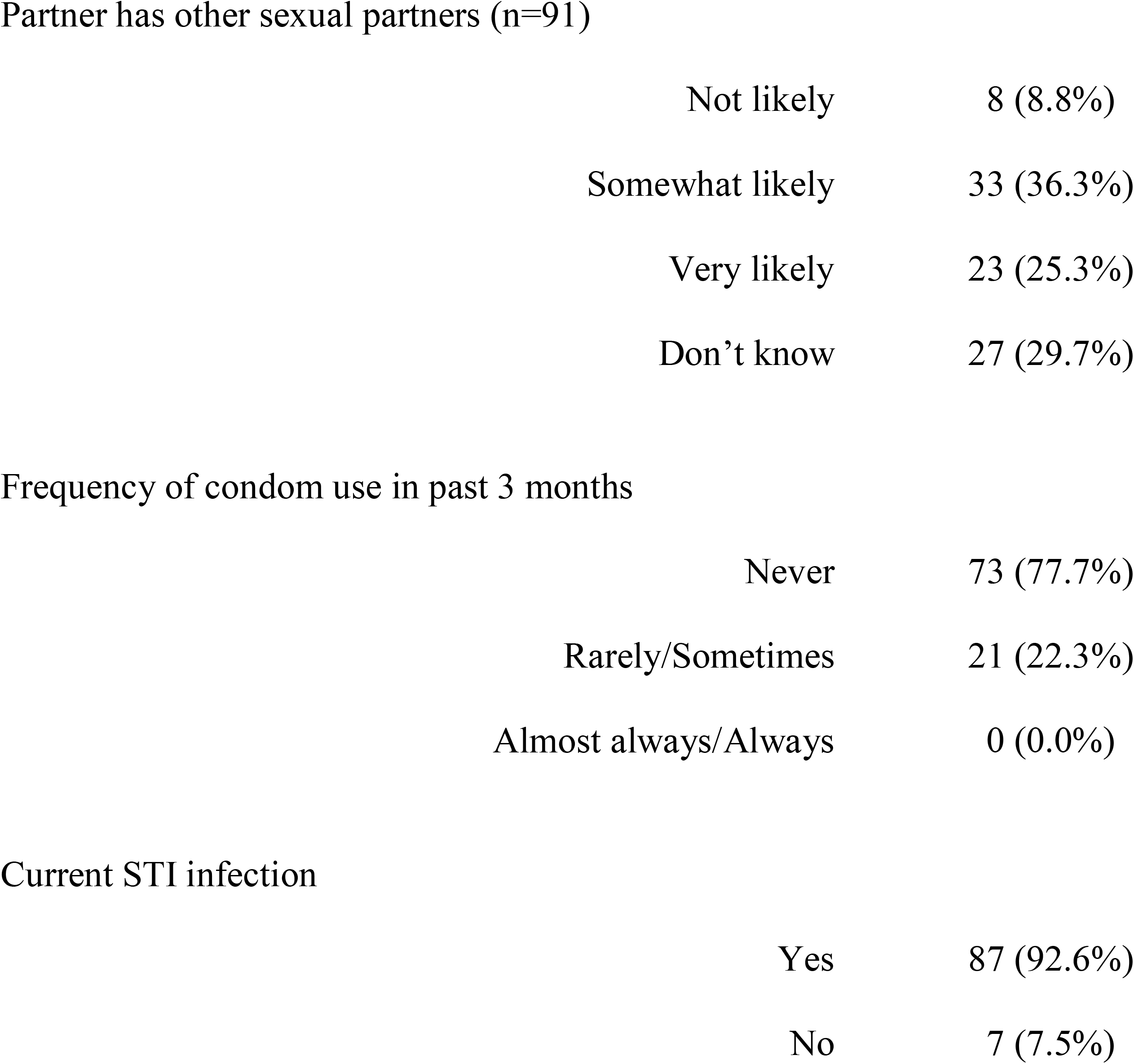
Sociodemographic and behavioral characteristics of women at high risk of HIV (using cutoff of 3+ risk factors) (n=94)

|  |  | N (%) |
| --- | --- | --- |
| Age |  |  |
|  | Mean (SD) | 25.8 (6.3) |
| Educational Attainment |  |  |
|  | Did not complete<br>secondary | 58 (61.7%) |
|  | Completed Secondary | 36 (38.3%) |
| Employment Status |  |  |
|  | Employed (full/part-time, formal/informal) | 28 (29.8%) |
|  | Student | 13 (13.8%) |
|  | Unemployed | 53 (56.4%) |
| Residence Type |  |  |
|  | Informal Dwelling | 58 (61.7%) |
|  | Formal Dwelling | 36 (38.3%) |
| Past year IPV |  |  |
|  | Yes | 13 (13.8%) |
|  | No | 81 (86.2%) |

|  |  |
| --- | --- |
| Yes | 55 (58.5%) |
| No | 39 (41.5%) |

|  |  |
| --- | --- |
| Yes | 8 (8.5%) |
| No | 86 (91.5%) |

Depression
|  |  |
| --- | --- |
| Depression (EPDS<br>≥11) | 5 (5.3%) |
| No Depression | 89 (94.7%) |

Pregnancy Timing
|  |  |
| --- | --- |
| Pregnancy happened at<br>the right time | 40 (42.6%) |
| Pregnancy happened at<br>the wrong time | 54 (57.5%) |

Feelings about pregnancy
|  |  |
| --- | --- |
| I wanted to have a baby | 30 (31.9%) |
| I have mixed feelings | 11 (11.7%) |
|  | about having a baby |  |
|  | I did not want to have a baby | 53 (56.4%) |
| Current Partner |  |  |
|  | No partner | 1 (1.1%) |
|  | Yes, the partner is my unborn baby's father | 91 (96.8%) |
|  | Yes, the partner is not my unborn baby's father | 2 (2.1%) |
| Relationship Status (n=93) |  |  |
|  | Cohabiting | 21 (22.6%) |
|  | Not Cohabiting | 72 (77.4%) |
| Primary Partner's HIV status |  |  |
|  | Positive | 5 (5.3%) |
|  | Negative | 2 (2.1%) |
|  | Don't know | 86 (91.5%) |
|  | No Recent Partner | 1 (1.1%) |
| Multiple partners in past 3 months |  |  |
|  | Yes | 14 (14.9%) |
|  | No | 80 (85.1%) |

Partner has other sexual partners (n=91)
|  |  |
| --- | --- |
| Not likely | 8 (8.8%) |
| Somewhat likely | 33 (36.3%) |
| Very likely | 23 (25.3%) |
| Don't know | 27 (29.7%) |

Frequency of condom use in past 3 months
|  |  |
| --- | --- |
| Never | 73 (77.7%) |
| Rarely/Sometimes | 21 (22.3%) |
| Almost always/Always | 0 (0.0%) |

Current STI infection
|  |  |
| --- | --- |
| Yes | 87 (92.6%) |
| No | 7 (7.5%) |

### Bivariate and multivariable regression analysis of alcohol use on HIV risk

Figure 1 presents the results from bivariate and multivariable analysis of the association between past year alcohol use prior to pregnancy and HIV risk. In multivariable analysis, the model was adjusted for the following potential confounders: age, education level, employment status, residence type and current partner. Using the cutoff of two or more risk factors to denote “high risk” for HIV acquisition, after adjusting for covariates, persons reporting past year alcohol use prior to pregnancy had 1.33 times greater odds of being in the high HIV risk category than persons reporting no alcohol use during that same time frame (adjusted OR 1.33, 95% CI 1.05-1.68). Using the cutoff of three or more risk factors to denote “high risk” after adjusting for these covariates, persons reporting past year alcohol use prior to pregnancy had 1.47 times greater odds of being in the high HIV risk category than persons reporting no alcohol use during that same time frame (adjusted OR 1.47, 95% CI 0.95-2.27).

**Figure 1.**
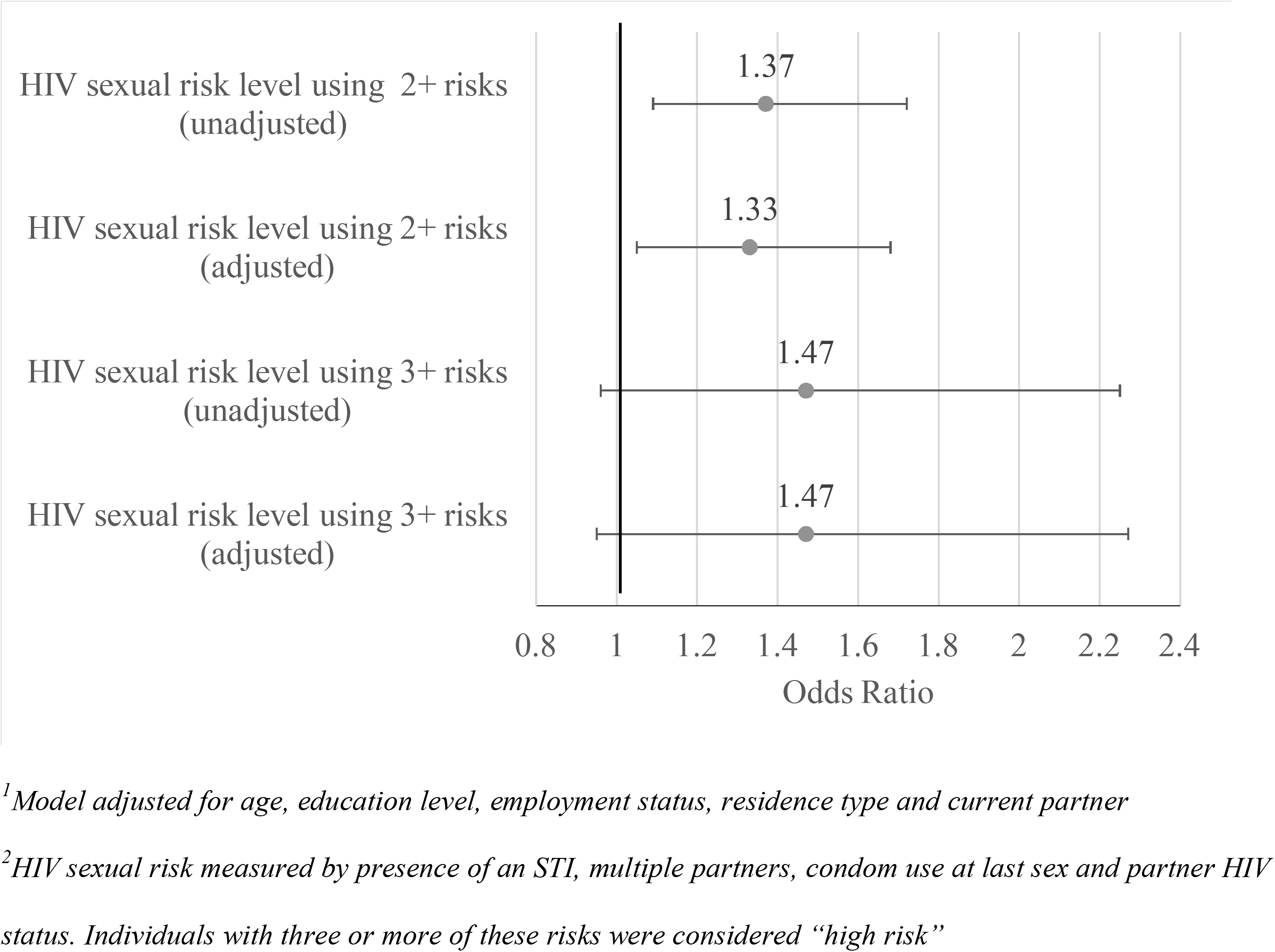
Unadjusted and adjusted^1^ analyses of associations between any alcohol use before pregnancy and HIV sexual risk

## Discussion

Using multiple validated measures of alcohol use and cutoffs consistent with the existing literature, the present study fills important gaps in the literature regarding the potential role alcohol may play as a risk factor for engaging in sexual risk behaviors in pregnancy by exploring associations between alcohol use and HIV sexual risk among HIV-uninfected pregnant women in South Africa. Obtaining accurate estimates of alcohol use during pregnancy presents challenges. Alcohol use during pregnancy is stigmatized and subject to underreporting due to social desirability bias (55). In our study sample, as expected the prevalence of alcohol use was much lower during pregnancy relative to prior to pregnancy (6% vs 50%). Nevertheless, the prevalence of alcohol use prior to pregnancy in our sample was higher than that observed in another recent study of HIV-uninfected women in Cape Town (50% vs 40%), but the prevalence of heavy alcohol use (defined as an AUDIT-C score ≥3) among those who reported any alcohol use, was similar in both samples at (66% vs 65%) (5). Observing a reduction of alcohol use and increased abstinence among pregnant women was expected; a large global body of evidence suggests that many women reduce their alcohol use or stop drinking entirely once their pregnancy is recognized (56–59). However, our estimate for alcohol use during pregnancy is lower than expected given estimates from prior studies. A recent systematic review and meta-analysis of alcohol use during pregnancy in sub-Saharan Africa found a pooled alcohol use during pregnancy prevalence of 24% (95% CI 22.48, 24.60) for the nine studies from South Africa (60). Underreporting of alcohol use during pregnancy in our sample may partially account for the low prevalence observed; timing of recognition of pregnancy may have also influenced this measure.

Women in our study who reported alcohol use prior to pregnancy differed in several ways from women who did not report this behavior. They had increased odds of reporting recent IPV, consistent with a prior study among women and men attending drinking establishments in Cape Town that found associations between IPV and both alcohol consumption and binge drinking among pregnant women (61). Pregnant women who drank alcohol also had increased odds of being depressed and reporting an unplanned pregnancy, consistent with findings from a meta-analysis where depression and unplanned pregnancy were both predictors of alcohol use during pregnancy (60). While it is known that pregnant women, including women in South Africa, experience stigma around alcohol use during pregnancy from healthcare providers (62–65), there is a dearth of literature exploring community norms and attitudes towards maternal drinking and how these may facilitate alcohol use in this population (63). Some qualitative work has been undertaken to explore pregnant women’s perceptions of these norms and attitudes (66), but there is a need for qualitative work with drinking partners, peers and family members to understand interpersonal and familial drivers of alcohol use during pregnancy and post-partum period.

Two recent systematic reviews of psychosocial interventions to address alcohol use in sub-Saharan Africa highlight the limited number of evidence-based interventions to address alcohol use in pregnancy in this setting (67, 68). The reviews found a total of two interventions (69–71) aimed at addressing alcohol use during pregnancy, both in South Africa. A brief intervention (BI) produced a significant reduction in mean AUDIT score immediately after intervention delivery but did not report outcomes in the post-partum period (71). A peer-led mentor mother intervention providing a maternal care package (which included HIV prevention content and a single session on alcohol use) found increased abstinence 5 years postpartum but did not improve abstinence relative to the control group during the critical pregnancy and breastfeeding periods (70). Neither intervention was designed to address alcohol use among persons experiencing alcohol dependence. Resources to address alcohol and other substance dependence in South Africa are limited and socioeconomic inequities in access to such services exist (72). The disease burden associated with alcohol use and HIV during pregnancy and the dearth of available interventions to address these issues underscore the urgent need for culturally acceptable evidence-based interventions to address alcohol use (including alcohol dependence) and HIV prevention among pregnant women in South Africa.

The women in our study are at high risk of HIV infection generally, due to the high prevalence of HIV in their community and increased risk of HIV acquisition during pregnancy (34). Using both cut-offs (2+ risks and 3+ risks) we observed an effect in the expected direction with women reporting alcohol use having greater odds of being in the high HIV sexual risk category. The four risk factors including, partner HIV serostatus, having >1 recent sex partners, condomless sex at last sex and the presence of an STI, present opportunities for intervention. While the prevalence of having an HIV positive partner was low (2%) more than one-fifth of the women in our study (22%) were not aware of their partner’s HIV serostatus. When looking at women in our high-risk group, 92% were not aware of their partner’s status. In the last decade, HIV self-testing (HIVST) (73) and couples HIV testing have emerged as promising interventions for reaching people that would otherwise be unlikely to access testing, such as male partners (74, 75).

The high prevalence of persons with partners of unknown HIV serostatus and high risk of HIV acquisition in pregnancy underscore the importance of integrating PrEP as a female controlled option for protection against HIV acquisition and onward transmission in this population. When taken correctly and consistently, daily oral PrEP (TDF/FTC) is a safe and highly effective method of HIV prevention (76–78). Further understanding of the relationship between alcohol use and PrEP initiation and adherence is needed to determine if alcohol is a barrier to optimal uptake of this HIV prevention measure in this population.

The prevalence of other STIs was also high in our study sample (29%). While the prevalence of STIs was slightly higher among persons who use alcohol, this difference was not statistically significant. However, the high prevalence throughout the study sample makes addressing STIs in this population a public health priority. To avoid reinfection in these women, it is important that they disclose their status to their partner and encourage their partner to receive treatment. Qualitative work with HIV positive pregnant women in Pretoria highlights barriers and facilitators to partner disclosure of STI results that pregnant women experience; many participants indicated that fear of IPV was a barrier to disclosure (79). To avoid re-infection, interventions that support partner disclosure, expedited partner therapy and offer support services for women experiencing IPV are needed in addition to STI testing services (80).

Our study had several limitations. Data were cross-sectional, precluding our ability to infer directionality or causality and all our alcohol use and HIV sexual risk variables (except for STI testing) were self-reported and subject to underreporting due to social desirability bias. Our study also had several strengths; we used globally validated alcohol measures in a large sample of HIV uninfected pregnant women to provide timely estimates of the prevalence of alcohol use and other HIV sexual risk factors in a population at high risk of HIV infection.

## Conclusion

We found a high prevalence of alcohol use prior to pregnancy in our study sample which may reflect high levels of alcohol use in early pregnancy. Furthermore, alcohol use was associated with being at high risk of HIV acquisition in HIV-uninfected pregnant women. Evidence-based interventions to address alcohol use and other HIV risk behaviors during pregnancy in South Africa are desperately needed. We identified several modifiable behaviors that could be targeted in future interventions including, improved measures to increase partner HIV testing and disclosure, and testing/counseling around STI results disclosure which could be integrated into ANC service delivery. Future studies should use objective measures of alcohol use as well as self-reported measures collected in real time to reduce bias from recall and social desirability. Finally, qualitative work is needed to understand individual and community level drivers of alcohol use among pregnant and breastfeeding women in this setting to develop a culturally tailored intervention to address these issues in this population.

## Data Availability

Data is available upon request from study investigators. Please for more information

